# Longitudinal decline in striatal dopamine transporter binding in Parkinson’s disease: Associations with apathy and anhedonia

**DOI:** 10.1101/2022.07.11.22277484

**Authors:** Harry Costello, Yumeya Yamamori, Suzanne Reeves, Anette Schrag, Robert Howard, Jonathan P. Roiser

## Abstract

**Background:** Motivational symptoms such as apathy and anhedonia are common in Parkinson’s disease, respond poorly to treatment, and have been hypothesised to share underlying neural mechanisms. Striatal dopaminergic dysfunction is considered central to motivational symptoms in Parkinson’s disease, but the association has never been examined longitudinally. We investigated whether the progression of dopaminergic neurodegeneration was associated with emergent apathy and anhedonia symptoms in Parkinson’s disease.

**Methods:** Longitudinal cohort study of 412 newly diagnosed Parkinson’s disease patients followed over five years as part of the Parkinson’s Progression Markers Initiative (PPMI) cohort. Apathy and anhedonia were measured using a composite score derived from relevant items of the 15-item geriatric depression scale (GDS-15) and part I of the MDS-Unified Parkinson’s Disease Rating Scale (MDS-UPDRS). Dopaminergic neurodegeneration was measured using repeated ioflupane [123-I] single photon emission computed tomography imaging of striatal dopamine transporters (DAT).

**Results:** Linear mixed-effects modelling across all contemporaneous data points identified a significant negative relationship between striatal DAT specific binding ratio(SBR) and apathy/anhedonia symptoms, which emerged as Parkinson’s disease progressed (interaction: β=-0.09, 95%CI[-0.15 -0.03], p=0.002). Appearance and subsequent worsening of apathy/anhedonia symptoms began on average two years after diagnosis and below a threshold striatal DAT SBR level. The interaction between striatal DAT SBR and time was specific to apathy/anhedonia symptoms, with no evidence of a similar interaction for general depressive symptoms from the GDS-15 (excluding apathy/anhedonia items) (β=-0.06, 95%CI[-0.13 0.01]) or motor symptoms indexed by the MDS-UPDRS part III (β=0.20, 95%CI[-0.25 0.65]).

**Conclusion:** The relationship between the progression of dopaminergic neurodegeneration and emergent apathy/anhedonia symptoms supports a central role for dopaminergic dysfunction in motivational symptoms in Parkinson’s disease. Striatal DAT imaging may be a useful indicator of apathy/anhedonia risk that could inform intervention strategies.

## Introduction

Characterised by deposition of α-synuclein in neurons and dopaminergic neuronal death, Parkinson’s disease (PD) is a model of dopamine dysfunction.^1^ Since its first description over 200 years ago, PD has been conceptualised as a movement disorder.^2,3^ However, it is now known that non-motor neuropsychiatric symptoms are common, and these have a greater negative impact on health-related quality of life than motor symptoms.^4,5^

Depression is a clinically and mechanistically heterogeneous syndrome which has led efforts to define subtypes based on symptom profile.^6^ Motivational symptoms including apathy and anhedonia contribute to an ‘interest-activity’ cluster of depressive symptoms that predicts worse response to antidepressants in non-PD depression.^7^ Apathy and anhedonia are particularly common in PD, estimated to affect 40%^8^ and 46%^9^ of patients, respectively. Both are syndromes of motivation: apathy is defined as diminished initiation of and engagement in activities; while anhedonia, though originally conceptualised solely as an inability to experience pleasure, is now recognised to incorporate a loss of interest (that is, motivation) to act in order to seek pleasure.^10^ This key motivational element is supported by evidence that depressed patients with marked anhedonia retain hedonic capacity.^11^ Treatments for apathy and anhedonia are limited, and the presence of either predicts greater illness severity and poorer quality of life in patients with both depression^12,13^ and PD.^14,15^

Recent research suggests that apathy and anhedonia overlap and share underlying neurobiological mechanisms related to reward processing.^10^ Human and animal studies have shown that dopaminergic transmission is crucial in reward processing, especially motivated behaviour^16^. Off dopaminergic medication, PD patients have greater reward processing deficits^17^ and report worse apathy^18^ and anhedonia^19^. Double-blind randomised controlled trials (RCT) of dopaminergic agonists have indicated potential therapeutic efficacy in treating apathy^20^ and depression^21^ in PD, though findings have been mixed^22^ and better powered trials are needed.

Consistent with a role for dopamine in motivational symptoms, the ventral striatum, which receives extensive dopaminergic innervation from the midbrain, has been implicated in apathy and anhedonia.^10^ Stroke lesions in the ventral striatum and caudate nucleus are associated with the development of apathy, while greater atrophy and lower metabolism in the ventral striatum have been associated with apathy in PD.^23,24^ Unmedicated depressed individuals exhibit attenuated striatal activation during reward processing,^25^ and a large longitudinal study of motivational processing in adolescents identified a robust inverse relationship between reward-related ventral striatal activation and both current and future anhedonia specifically, over and above low mood.^26^

Dopamine transporter (DAT) imaging is commonly used as a diagnostic tool in PD, as it is sensitive to degeneration of dopaminergic nigrostriatal pathways.^27^ Striatal DAT decreases in PD as the disease progresses owing to a loss of presynaptic dopaminergic projections from the substantia nigra and ventral tegmental area. DAT imaging, using Ioflupane [123-I] single photon emission computed tomography (SPECT), is clinically useful in distinguishing neurodegenerative Parkinson’s syndromes from cases of essential tremor or drug-induced parkinsonism,^28^ but is a poor predictor of the progression of motor symptoms.^29^ Animal studies have shown that selective lesions of substantia nigra dopamine neurons can induce motivational deficits and affective impairments, without eliciting motor deficits. In PD patients, some small cross-sectional studies have reported that apathy and depression are associated with lower striatal DAT binding, although findings are mixed.^22,30,31^ However, no study to date has examined the longitudinal relationship between striatal DAT binding and apathy or anhedonia.

Therefore, we investigated the relationship between dopaminergic degeneration and the emergence and progression of apathy and anhedonia in PD longitudinally. We used data from the Parkinson’s Progression Markers Initiative (PPMI) cohort, an international multicentre cohort study. We tested the hypothesis that apathy and anhedonia symptoms in PD are driven by dopaminergic degeneration, indexed by DAT binding in the striatum. We predicted a negative relationship between striatal DAT binding and apathy and anhedonia, and that any such relationship would be specific to these symptoms, and not observed with general depressive symptoms.

## Methods

### Participants

All data were obtained from the PPMI database (https://www.ppmi-info.org/), first accessed July 2021. Launched in 2010 to identify markers of Parkinson’s onset and progression, PPMI includes repeated clinical measures and imaging biomarkers including [123-I]-SPECT.

PPMI enrolled untreated, newly diagnosed Parkinson’s patients and age- and sex-matched healthy controls. All participants underwent a standard battery of assessments^32^ including the MDS-Unified Parkinson’s Disease Rating Scale (MDS-UPDRS), the 15-item Geriatric Depression Scale (GDS) and DAT imaging with [123I]-SPECT repeatedly, over five years.

The current analysis included participants with a diagnosis of PD at baseline. In order to avoid capturing chronic symptoms in the context of pre-morbid depression prior to the development of PD, all participants who had received a diagnosis of major depressive disorder more than five years prior to diagnosis with PD were excluded (N=11).

### Apathy and anhedonia measure

We created a composite measure of apathy and anhedonia comprising items from the MDS-UPDRS and GDS-15 (Table 1). The apathy/anhedonia score was operationalised based on the three-item ‘apathy/anhedonia’ factor of the GDS-15, which has been previously validated in older adults,^33^ and the apathy-specific measure in part I of the MDS-UPDRS.^34^

**Table 1.** Apathy/anhedonia composite measure.

| **Table 1. Apathy/anhedonia composite measure** |  |
| --- | --- |
| **MDS-UPDRS part I, item 1.5:** | Over the past week, have you felt indifferent to doing activities or being with people?<br>Scale 0-4 (normal – severe) |
| **GDS item 2:** | Have you dropped many of your activities or interests?<br>Scale yes (1) / no (0) |
| **GDS item 9:** | Do you prefer to stay at home, rather than going out and doing new things?<br>Scale yes (1) / no (0) |
| **GDS item 13:** | Do you feel full of energy?<br>Scale yes (0) / no (1) |

A nine-item “depression” factor from the GDS-15 (excluding the above apathy/anhedonia items) was used as a comparative measure to test whether any relationship with striatal DAT binding was specific to apathy/anhedonia.^35^ This “depression” factor was identified in an independent cohort using factor analysis to investigate depressive symptom clusters in patients with PD, and comprises GDS-15 items 1, 3, 5, 7-8, 11-12, and 14-15.^35^

### Image processing and calculation of striatal DAT specific binding ratio (SBR) by PPMI

All images were analysed according to the PPMI imaging technical operations manual and had undergone analysis to determine striatal DAT specific binding ratio (SBR) (see Supplementary Methods for details).^36^ Regions of interest (ROI) included the left and right caudate, the left and right putamen, and the occipital cortex (reference tissue). Count densities for each region were extracted and used to calculate the SBR for each of the striatal ROI. SBR is calculated as: (target region/reference region)–1. To account for asymmetry, the minimum SBR for each region was used.

#### Statistical analysis

We used linear mixed-effects modelling to examine the relationship between apathy/anhedonia score (dependent variable) and striatal DAT SBR (independent variable), which were acquired contemporaneously, and how this relationship changed over the progression of illness. This involved fitting both the main effect of striatal DAT SBR (across all time-points) and its interaction with time. This allowed inter-individual heterogeneity and unequal follow-up intervals to be accommodated by incorporating random effects. Random intercept terms at the participant level were tested, with random slopes for time (defined as year of follow-up assessment). The interaction term between striatal DAT SBR and time allowed us to assess how the relationship between apathy/anhedonia and striatal DAT SBR changed over time, using all available data. When a significant interaction was identified, post-hoc tests were conducted using Pearson’s correlations at each timepoint separately.

Two sets of regression were conducted: 1) unadjusted and 2) adjusted for age, sex, years of education, number of missing years and duration of PD (all at baseline); plus cognition (Montreal Cognitive Assessment), severity of motor symptoms (MDS-UPDRS part III score, ‘off’ medication), stage of disease/functional disability (Hoehn and Yahr scale), levodopa equivalent dose (LED), antidepressant medication status and the GDS-15 “depression” factor (all at each contemporaneous timepoint). Model fit was tested using the Akaike information criterion (AIC).

To test whether findings were specific to apathy/anhedonia, mixed-effects modelling was repeated with motor symptom severity and the GDS-15 ‘depression’ factor as dependent variables in separate analyses, incorporating apathy/anhedonia score as a covariate.

The “two-lines” test (see Supplementary Methods) was performed to test the validity of a threshold effect of striatal DAT SBR on apathy/anhedonia.^37^ The two-lines test, which relies on using two regression lines (one for low values of the predictor, the other for high values), has been proposed as a more effective and valid method than quadratic regression modelling to indicate a non-linear relationship. Using this method also allows identification of a “breakpoint” value, using the “Robin Hood” algorithm. The breakpoint value is identified by reallocating observations from the statistically stronger of the two lines to the weaker, increasing statistical power to detect the inverse unimodal shape. For an association to qualify as a U-shaped using the two-lines test, the two regression lines must differ in direction in addition to being independently significant.^37^ However, it should be noted that the threshold value identified by the Robin Hood algorithm is not directly interpretable, as the break point is identified from a range of candidate values, meaning that it is not estimated precisely.

All statistical analyses were performed in R version 4.1.2. The R package ‘lme4’ was used for mixed effects modelling, and the ‘twolines’ package was used for the two-lines test.

## Results

### Participant characteristics

A total of 412 participants with PD were included at baseline with dropout of one-quarter of participants by year five (Table 2).

**Table 2.** Characteristics of PPMI participants with Parkinson’s disease.

|  |  |  |  |  |  |  |
| --- | --- | --- | --- | --- | --- | --- |
|  | **Baseline** | **Year 1** | **Year 2** | **Year 3** | **Year 4** | **Year 5** |
| **n** | 412 | 384 | 367 | 355 | 335 | 304 |
| **Age at entry** | 61.77<br>(9.76) | 61.66<br>(9.86) | 61.66<br>(9.86) | 61.64<br>(9.86) | 61.40<br>(9.96) | 60.94<br>(9.84) |
| **Age at diagnosis** | 61.22<br>(9.73) | 61.1<br>(9.83) | 61.1<br>(9.84) | 61.09<br>(9.82) | 60.86<br>(9.92) | 60.39<br>(9.83) |
| **Duration of PD at cohort entry (years)** | 6.56 (6.39) | 6.64 (6.51) | 6.70 (6.56) | 6.65 (6.54) | 6.57 (6.48) | 6.6 (6.53) |
| **Years of education** | 15.54<br>(2.99) | 15.47<br>(2.9) | 15.6<br>(2.87) | 15.61<br>(2.92) | 15.62<br>(2.90) | 15.61<br>(2.97) |
| **Male (%)** | 273 / 412<br>(66%) | 255 / 384<br>(66%) | 244 / 367<br>(66%) | 235 / 355<br>(66%) | 225 / 335<br>(67%) | 205 / 304<br>(67%) |
| **Apathy/anhedonia composite** | 1.21 (1.14) | 1.41 (1.30) | 1.51 (1.43) | 1.48 (1.48) | 1.58 (1.49) | 1.67 (1.54) |
| **MDS-UPDRS III** | 20.93<br>(8.80) | 25.25<br>(10.92) | 27.62<br>(11.39) | 29.58<br>(12.35) | 30.97<br>(12.20) | 31.27<br>(12.28) |
| **UPDRS total** | 32.41<br>(13.14) | 39.59<br>(16.12) | 43.02<br>(16.97) | 46.33<br>(18.80) | 48.70<br>(19.68) | 49.96<br>(19.04) |
| **MOCA** | 27.13<br>(2.28) | 26.28<br>(2.82) | 26.27<br>(3.14) | 26.37 (3.01) | 26.42 (3.57) | 26.53 (3.53) |
| **GDS** | 2.29 (2.43) | 2.53 (2.92) | 2.58 (2.87) | 2.58 (2.79) | 2.59 (2.84) | 2.78 (2.80) |
| **GDS >5** | 55 / 412<br>(13%) | 61 / 384<br>(16%) | 63 / 366<br>(17%) | 58 / 355<br>(16%) | 58 / 333<br>(17%) | 61 / 303<br>(20%) |
| **STAI** | 65.05<br>(18.14) | 64.92<br>(18.63) | 64.65<br>(18.33) | 64.41<br>(18.38) | 64.71<br>(18.71) | 64.60<br>(18.95) |
| **% commenced PD medication** | 0/412 (0%) | 225/382<br>(59%) | 310/367<br>(84%) | 329 (355)<br>92.7% | 319/332<br>(96.1%) | 293/304<br>(96.4%) |
| **% on antidepressant** | 29/412<br>(7.0%) | 29/384<br>(7.6%) | 33/367<br>(9.0%) | 30/355<br>(8.5%) | 32/335<br>(9.6%) | 26/278<br>(9.4%) |
| **No. DAT scans** | 408 | 360 | 341 | 10 | 280 | 7 |
| **Striatal DAT SBR** | 2.50 (0.75) | 2.21 (0.66) | 2.07 (0.68) | 1.67 (0.54) | 1.82 (0.64) | 1.30 (0.67) |
| Mean (SD); N (%); MDS-Unified Parkinson's Disease Rating Scale (UPDRS); Montreal Cognitive Assessment (MOCA); Geriatric Depression Scale (GDS); State-Trait Anxiety Inventory (STAI); dopamine transporter specific binding ratio (DAT SBR) |  |  |  |  |  |  |

Almost all (98.8%) DAT imaging was completed at baseline and in years one, two and four, with only 17 participants imaged in years three and five. However, due to the mixed-effects modelling approach we could incorporate all available data into the analysis. As expected, at baseline striatal DAT SBR in PD participants was on average around half that of healthy controls; and there was evidence of a marked decline over time (mean±SD percentage reduction from baseline: year 1=-9.7±17.4%, year 2=-16.6±17.7%, year 4=-26.6±18.4%; Figure 1A).

**Figure 1.**
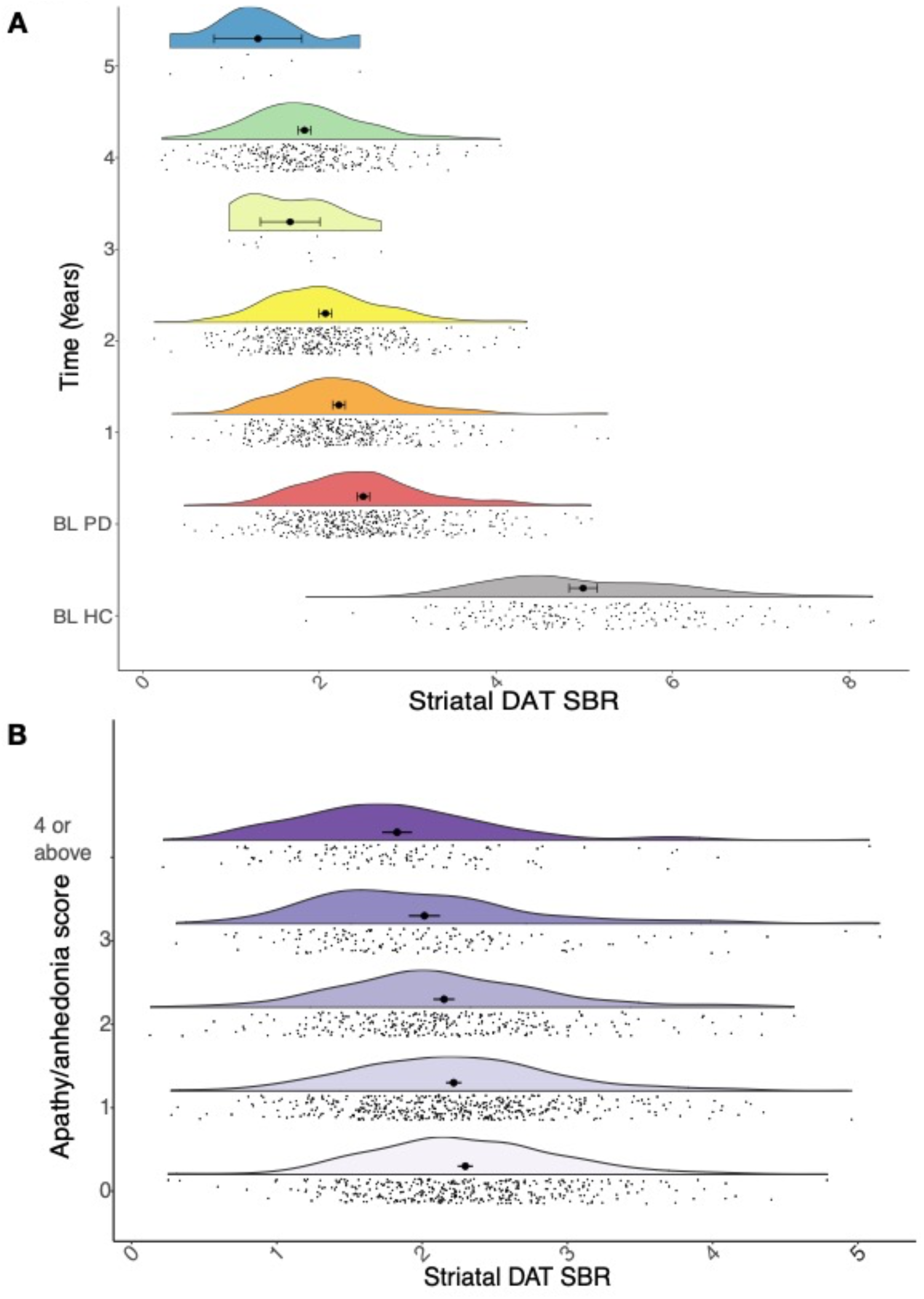
**A**. Raincloud plots showing the expected progressive reduction in striatal dopamine transporter (DAT) specific binding ratio (SBR) in Parkinson’s disease (PD, coloured plots) over time relative to baseline (BL), compared to healthy controls (HC, grey plot) at BL. **B**. Raincloud plots showing striatal DAT SBR at different levels of apathy/anhedonia, across all years.

As anticipated, motor symptom severity increased over time (MDS-UPDRS III: β=4.0, 95%CI [3.7 4.3], p<0.001). Although all participants were unmedicated at baseline, a majority (59%) had commenced dopaminergic medication by year one. One-quarter of participants (25%) with PD were taking antidepressant medication at baseline, but only 13% had a GDS-15 score >5 (suggestive of clinical depression) and indication for treatment was not available.

Parallel factor analysis of the apathy/anhedonia composite measure indicated a single factor loading with acceptable internal consistency (Cronbach’s alpha=0.60, 95%CI [0.56 0.62]). Apathy/anhedonia composite score increased over time (β=0.22, 95%CI [0.10 0.34], p<0.001) (Figure 1B).

### Relationship between striatal DAT SBR and apathy/anhedonia

Longitudinal analysis revealed that, while the overall relationship between striatal DAT SBR and apathy/anhedonia across all timepoints was non-significant (β=0.09, 95%CI [-0.06 0.24], p=0.2), there was a significant interaction with time in the unadjusted model (β=-0.09, 95%CI [-0.15 -0.03], p<0.001) (Figure 2,3A). A similar relationship was observed in the adjusted model (including: demographic factors; cognition; motor symptoms; LED; antidepressant status; and the GDS-15 “depression” factor, which excludes apathy/anhedonia items), which was more parsimonious (AIC: adjusted=3143.5, unadjusted=3762.1). Post-hoc analysis revealed that striatal DAT SBR was not significantly associated with apathy/anhedonia at baseline (r=0.03, p=0.54), but this relationship emerged over follow up (year 4: r=-0.26, p<0.01), strengthening as time progressed. In other words, as striatal DAT SBR decreases and apathy/anhedonia increases over the course of disease progression, a negative relationship between the two develops (Figure 3B). Similar results were observed in separate analyses of the caudate and putamen striatal subdivisions (Supplementary Results).

**Figure 2.**
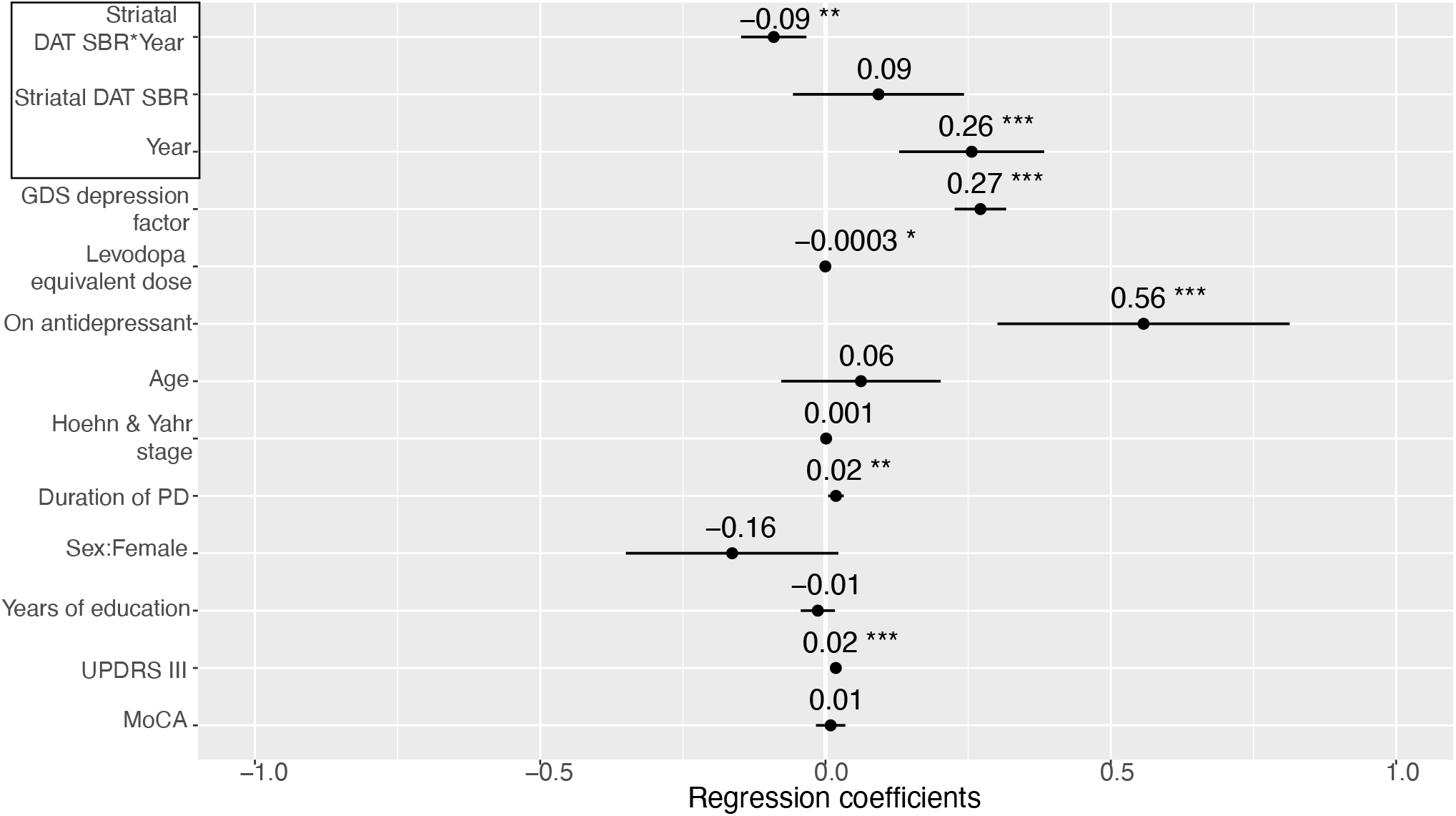
Mixed-effects model investigating the relationship between striatal dopamine transporter (DAT) specific binding ratio (SBR) and apathy/anhedonia longitudinally, adjusted for age, sex, years of education, and duration of Parkinson’s disease (PD) (assessed at baseline only); as well as cognition (assessed with the Montreal Cognitive Assessment (MoCA)), severity of motor symptoms (MDS-UPDRS part III score), functional disability (Hoehn & Yahr scale), levodopa equivalent dose, antidepressant status, and the Geriatric Depression Scale-15 (GDS) “depression” factor (assessed contemporaneously). Points represent estimated regression coefficients and bars represent 95% confidence intervals; p<0.05*, p<0.01**, p<0.001***.

**Figure 3.**
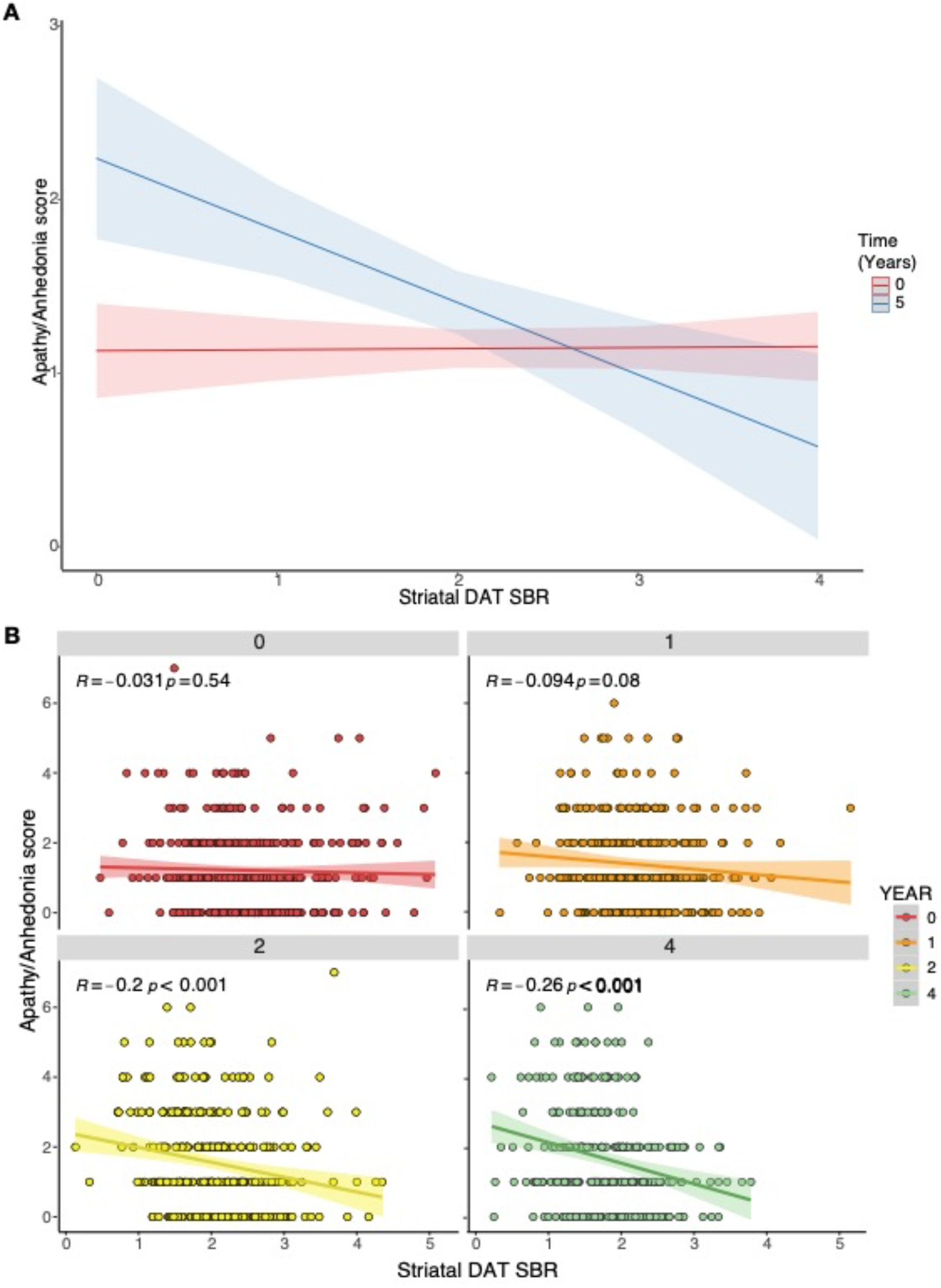
**A**. Adjusted mixed-effects model simulation of predicted apathy/anhedonia score by striatal dopamine transporter (DAT) specific binding ratio (SBR) and time (years 0 & 5 simulated). **B**. Scatter plots displaying unadjusted linear regressions between striatal DAT SBR and apathy/anhedonia score at years 0 (baseline), 1, 2 and 4, showing the strengthening of the relationship over time.

This interaction between striatal DAT SBR and time was only observed for apathy/anhedonia. Separate sensitivity analyses using MDS-UPDRS part III score (Figure S1A) and the GDS-15 “depression” factor (Figure S1B) as dependent variables, using an adjusted model including apathy/anhedonia as a covariate, showed no significant interactions with time. Though motor symptoms increased over time and lower striatal DAT SBR was associated with worse motor symptoms across all timepoints (main effect: β=-1.30, 95%CI [-2.4 -0.22], p=0.02) (Figure S1A), unlike apathy/anhedonia, this association did not strengthen over time (interaction: β=0.20, 95%CI [-0.25 0.65], p=0.4) (Figure S1A). We found no significant relationship between striatal DAT SBR and the GDS-15 “depression” factor (main effect across all timepoints: β=-0.06, 95%CI [-0.14 0.26], p=0.5; interaction with time: β=-0.06, 95%CI [-0.13 0.01], p=0.09) (Figure S1B).

To assess the clinical relevance of the association between apathy/anhedonia and striatal DAT SBR we analysed the three items of the GDS contributing to our apathy/anhedonia measure as the dependent variable, which has previously been validated as a clinical measure of apathy (GDS-3A).^33^ A GDS-3A score of ≥2 has high specificity for clinically relevant apathy in older adults, equivalent to a score of ≥14 using the Starkstein apathy scale.^33^ In adjusted analyses, similar results were observed for both continuous and categorical analyses of the GDS-3A, with significant interactions between striatal DAT SBR and time (Figure S3A&B).

### Is the association between striatal DAT SBR and apathy/anhedonia dependent on change from baseline?

Restricting analysis to post-baseline timepoints, we incorporated baseline DAT SBR as a covariate within the adjusted model (Figure S2), and also examined change-from-baseline DAT SBR, in separate analyses (Supplementary Results). There was no evidence that baseline DAT SBR predicted later apathy/anhedonia, and no evidence for an association with change-from-baseline DAT SBR (Figure 4, top); this is in contrast to the clear strengthening of association between absolute striatal DAT SBR and apathy/anhedonia score as PD progresses (Figure 4, bottom).

**Figure 4.**
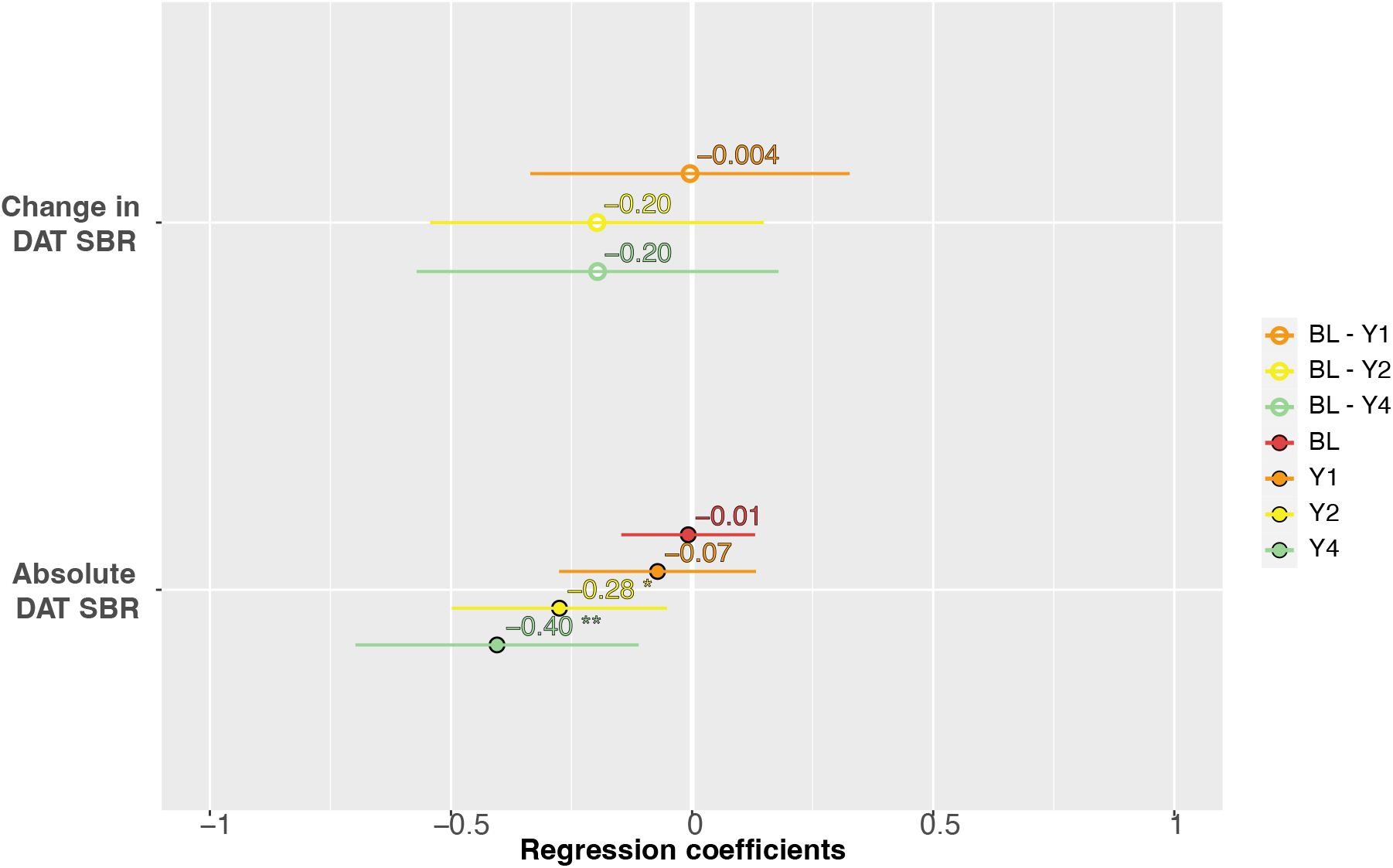
Association between striatal dopamine transporter (DAT) specific binding ratio (SBR) and apathy/anhedonia score using change-from-baseline (BL) SBR values (top) and absolute SBR values (bottom). Individual linear regression models at different years are displayed. All change-from-baseline models were non-significant. Points represent estimated regression coefficients and bars represent 95% confidence intervals; p<0.05*, p<0.01**, p<0.001***.

These analyses suggest that it is the absolute value of striatal DAT SBR, and not change from baseline, that is associated with apathy/anhedonia.

### Exploratory analysis of threshold striatal DAT SBR in developing apathy/anhedonia

Using the two-lines test^37^ we assessed whether a threshold striatal DAT SBR value exists, below which the relationship with apathy/anhedonia begins to emerge.

Analysis including all available contemporaneously acquired datapoints, across all years, revealed the threshold effect, consistent with a non-linear relationship (Figure 5). Both the upward (p=0.03) and downward (p<0.001) regression slopes achieved statistical significance (Figure 5). A threshold striatal DAT SBR value of ∼2.67 was estimated, below which a negative relationship with apathy and anhedonia develops. This is consistent with an explanation according to which apathy/anhedonia symptoms start to increase beyond a certain level of dopaminergic degeneration. However, we note that the Robin-Hood algorithm used in the two-lines test primarily tests for the presence of a threshold and cannot precisely identify the precise turning point value. Therefore the significant upward regression slope is conceivably an artefact of the operation of the algorithm, rather than evidence of patients with the highest striatal SAT SBR having higher apathy/anhedonia scores, which was not evident in other analyses (Figure 3B).

**Figure 5.**
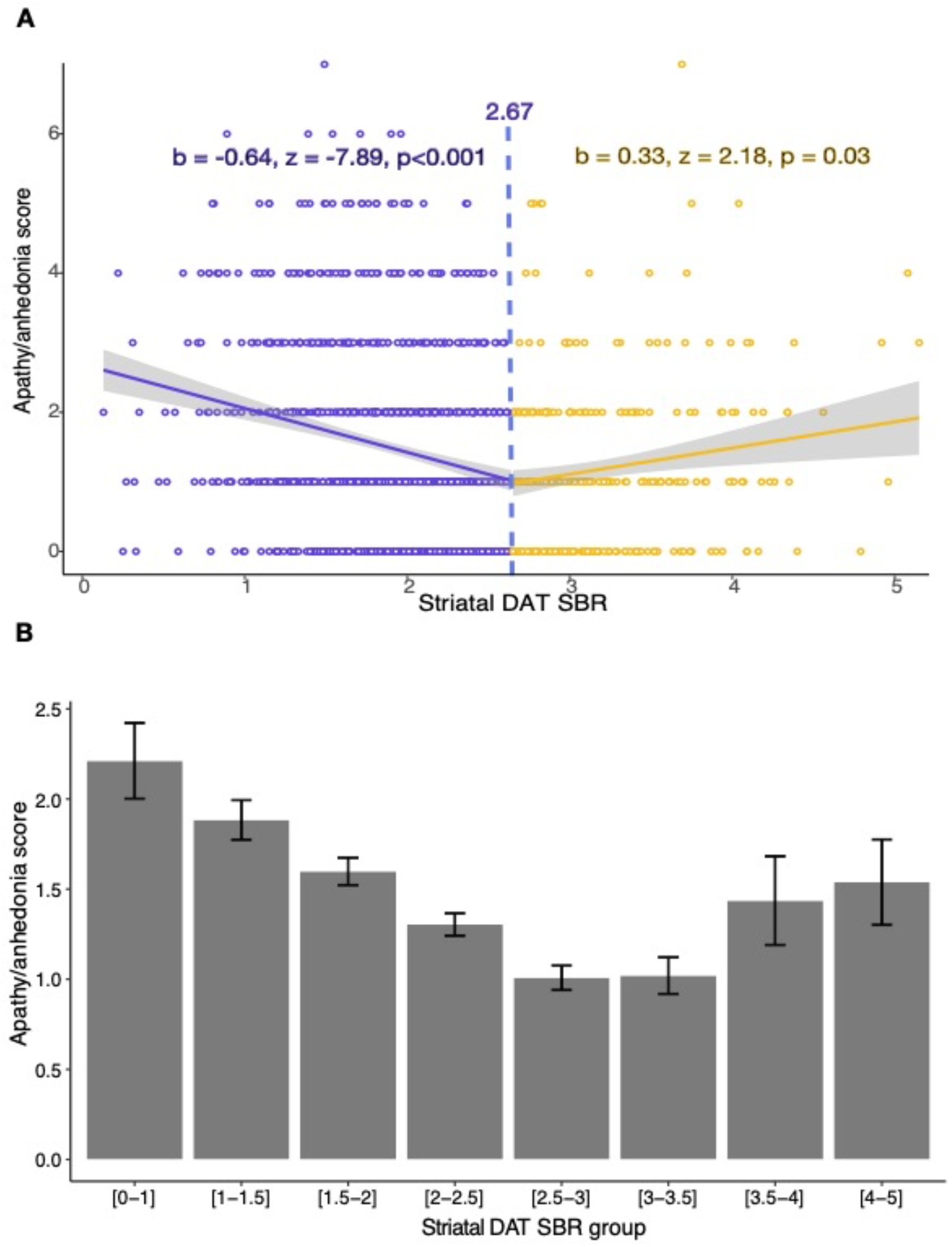
A. Two-lines test assessing the validity of a threshold effect between striatal dopamine transporter (DAT) specific binding ratio (SBR) and apathy/anhedonia score, across all available contemporaneously acquired datapoints and years of follow up. The two-lines test estimates a DAT SBR threshold of ∼2.7, beyond which apathy/anhedonia starts to increase. B. Mean apathy/anhedonia score and standard error at binned increments of SBR, across all available contemporaneously acquired datapoints and years of follow up.

## Discussion

This is the first longitudinal analysis of the relationship between striatal dopaminergic dysfunction and apathy and anhedonia in PD. Using a common clinically used neuroimaging marker, we found that a negative relationship between striatal DAT SBR and apathy/anhedonia symptoms emerged beyond a certain threshold of dopaminergic neurodegeneration. This relationship is not present during the early stages of illness, but develops over time as PD pathology progresses and a threshold level of pre-synaptic striatal dopaminergic neurodegeneration is crossed.

Our findings support existing evidence for the important role striatal dopamine dysfunction plays in disorders of motivation^10,23^ and suggests that striatal DAT imaging could contribute to indicating the risk of developing motivational symptoms in PD. In patients whose striatal DAT SBR is particularly low, there is scope to monitor apathy and anhedonia symptoms, or to intervene early with interventions such as behavioural activation therapy which has been shown to be potentially effective in treating these symptoms in PD.^38^ However, further validation of striatal DAT SBR as a potential marker of apathy/anhedonia risk is needed, especially as the Robin-Hood algorithm used in the two-lines test primarily tests for the presence of a threshold effect and cannot precisely identify the true turning point value.

We also caution that the significant upward regression slope identified by the two-lines test is conceivably an artefact of the algorithm, rather than evidence that patients with the highest striatal DAT SBR binding have higher apathy/anhedonia scores, as this was not consistent with other analyses. That said, we note that some prior studies have reported abnormally *high* DAT binding in depressed PD patients^39,40^, which could be interpreted as reflecting greater dopamine clearance via DAT, which would lead to lower striatal dopamine transmission. This could possibly represent an alternative mechanism driving apathy and anhedonia in PD patients, especially early in the disease course when dopamine neurodegeneration is less pronounced. However, this interpretation is speculative and needs to be tested in future studies.

In contrast to the pattern we observed in relation to apathy and anhedonia, striatal DAT SBR showed a consistent relationship with motor symptoms over time, with no evidence for an interaction. This aligns with previous studies,^29^ and may reflect differential degeneration of functional subdivisions of the striatum. At diagnosis with PD, it is likely that up to 80% of dopaminergic projections to the caudate have already been lost.^41,42^ Dopaminergic denervation of the caudal-motor subregion of the striatum, which receives input from the primary motor and premotor cortices, is central to the development of motor symptoms.^43^ Due to the degree of early denervation in this region, a floor effect is likely to explain why the relationship between striatal DAT SBR and motor symptoms does not emerge over time following diagnosis.

The association between striatal DAT SBR and apathy/anhedonia symptoms that emerges over time was present in putamen and caudate striatal subregions; however, the PPMI dataset does not include ventral striatum DAT binding. The ventral striatum, which receives dopaminergic input from the ventral tegmental area, is relatively spared in early Parkinson’s disease^42,43^ and is believed to play a crucial role in motivated behaviour. Atrophy in this region has been linked with apathy across neurodegenerative disorders.^23^ We speculate that the interaction we observed with time may occur as a consequence of the later degeneration and dopaminergic denervation of the ventral striatum, with motivational symptoms being less affected by the earlier caudal-motor striatal degeneration.

It is well established that dopamine’s effect on motivation reflects neuromodulation of fronto-striatal circuitry.^44,45^ The function of dopamine in the striatum has been proposed to depend on phasic and tonic dopaminergic cell firing modes.^46,47^ Short-latency phasic firing of dopaminergic neurons in the striatum encodes reward prediction errors, crucial for reinforcement learning, while tonic levels of activity are thought to signal average reward valuation.^44^ Animal studies have shown that different striatal regions receive distinct dopamine signals encoding different aspects of motivational stimuli and their prediction.^48^

A gradient in the rate of re-uptake of dopamine, from ventral to dorsal regions of the striatum, has been reported.^49^ This is consistent with the notion that that dorsal striatal regions are more sensitive to phasic signalling while the ventral striatum has greater utilisation of tonic signalling, which has been proposed to represent reward valuation.^44,49^ The pattern and timing of neurodegeneration in PD, which progresses from dorsal to ventral striatum, may explain why apathy/anhedonia symptoms occur later in disease, and supports existing evidence that anhedonia and apathy are a consequence of how reward valuation is represented by dopaminergic signalling.^50^

Though our findings are specific to PD, they may also reveal mechanistic insights into apathy and anhedonia transdiagnostically. Individual variation in the basal tone of different dopaminergic projection systems is hypothesized to be crucial in cognitive biases and susceptibility to psychiatric symptoms such as apathy and anhedonia.^44^ However, DAT imaging in depression has been conducted only in small samples, with conflicting findings.^51^ Future studies are required to further explore the relationship between striatal dopaminergic dysfunction and the emergence of specific neuropsychiatric symptoms. Additionally, the role of dopaminergic medication in the treatment of apathy and anhedonia requires further investigation. Methylphenidate, a noradrenaline-dopamine reuptake inhibitor, has recently been found to have efficacy in treatment of apathy in Alzheimer’s dementia.^52^ Further understanding of the effects of dopaminergic medication regime on apathy and anhedonia in PD is needed, in addition to RCTs of dopaminergic agents for apathy and anhedonia across different disorders.

## Limitations

The PPMI cohort also only includes recently diagnosed patients with PD, and may not be applicable to individuals in the later stages of the condition where the spread of neurodegeneration and systems involved are likely more complex.

Though our composite measure of apathy and anhedonia exclusively incorporated items specifically designed to measure these symptoms, it has not been validated previously, and it is unclear from this measure alone whether participants developed clinically meaningful levels of apathy and anhedonia during the study. However, we addressed this by additionally examining a measure that has been clinically validated (the GDS-3A^33^, which formed the bulk of our apathy/anhedonia measure). This showed results consistent with our primary analyses, with clear interactions with time using both continuous and categorical apathy measures based. However, we note that the GDS-3A has low sensitivity for apathy, meaning clinically relevant cases will be missed by this measure. It would have been preferable to have used validated measures of apathy and anhedonia, and to have assessed whether these symptoms were clinically relevant, but unfortunately this information is not available in the PPMI dataset.

The associations we observed with between apathy and anhedonia and striatal DAT SBR potentially could be a consequence of other Parkinson’s symptoms, functional disability or depression. However, this is unlikely as all models were adjusted for cognition, motor symptoms, disease duration, functional disability, and general depressive symptoms. Additionally, our findings were relatively specific to apathy/anhedonia with no significant relationship observed between striatal DAT SBR and general depressive symptoms in sensitivity analyses; although we note that the upper end of the 95% confidence interval for general depression scores was near zero, and therefore it is possible that a weak interaction exists which we did not have statistical power to detect, even in the large PPMI dataset.

Although DAT imaging is used clinically as a measure of dopaminergic neurodegeneration in PD, the radioligand [123I]-Ioflupane used in the PPMI cohort has also been suggested to have a modest affinity for the serotonin transporter.^53^ The degree to which DAT imaging reflects solely dopaminergic function is therefore uncertain. However, it is known that individual neurons can release multiple transmitters from the same synapse^54^ and recent research suggests co-release of dopamine and serotonin is functionally relevant,^55^ although co-localisation of different neurotransmitter transporters is sparse.^56^ As a consequence it is likely that the interplay between dopamine and serotonin in the striatum is important in motivational symptoms, as suggested by animal studies.^48,50^ LED and antidepressant medication status were also incorporated into our modelling, with no effect on the interaction between DAT SBR and time.

Finally, although healthy controls underwent SPECT imaging at baseline and had average striatal DAT SBR levels almost double that of Parkinson’s participants, they did not have repeat imaging, so comparison to a control group longitudinally was not possible.

## Conclusion

In PD, striatal DAT SBR is associated with apathy and anhedonia symptoms over time. As dopaminergic neurodegeneration progresses and striatal DAT SBR declines, a threshold is reached beyond which apathy and anhedonia symptom burden begins to increase. This effect is relatively specific to apathy and anhedonia, with no such interaction evident for depressive or motor symptoms, and is consistent with prior evidence of the crucial role of striatal dopaminergic dysfunction in motivational dysfunction. Further validation of striatal DAT imaging as a potential clinically useful marker of apathy and anhedonia risk in Parkinson’s disease is warranted.

## Data Availability

The PPMI cohort is an existing longitudinal cohort & open dataset. All data from this study is available upon request.

## Acknowledgements

We thank the Michael J. Fox Foundation, and the investigators, Parkinson’s patients and controls that enabled the Parkinson’s Progression Marker Initiative.

## Supplement

**Figure S1.**
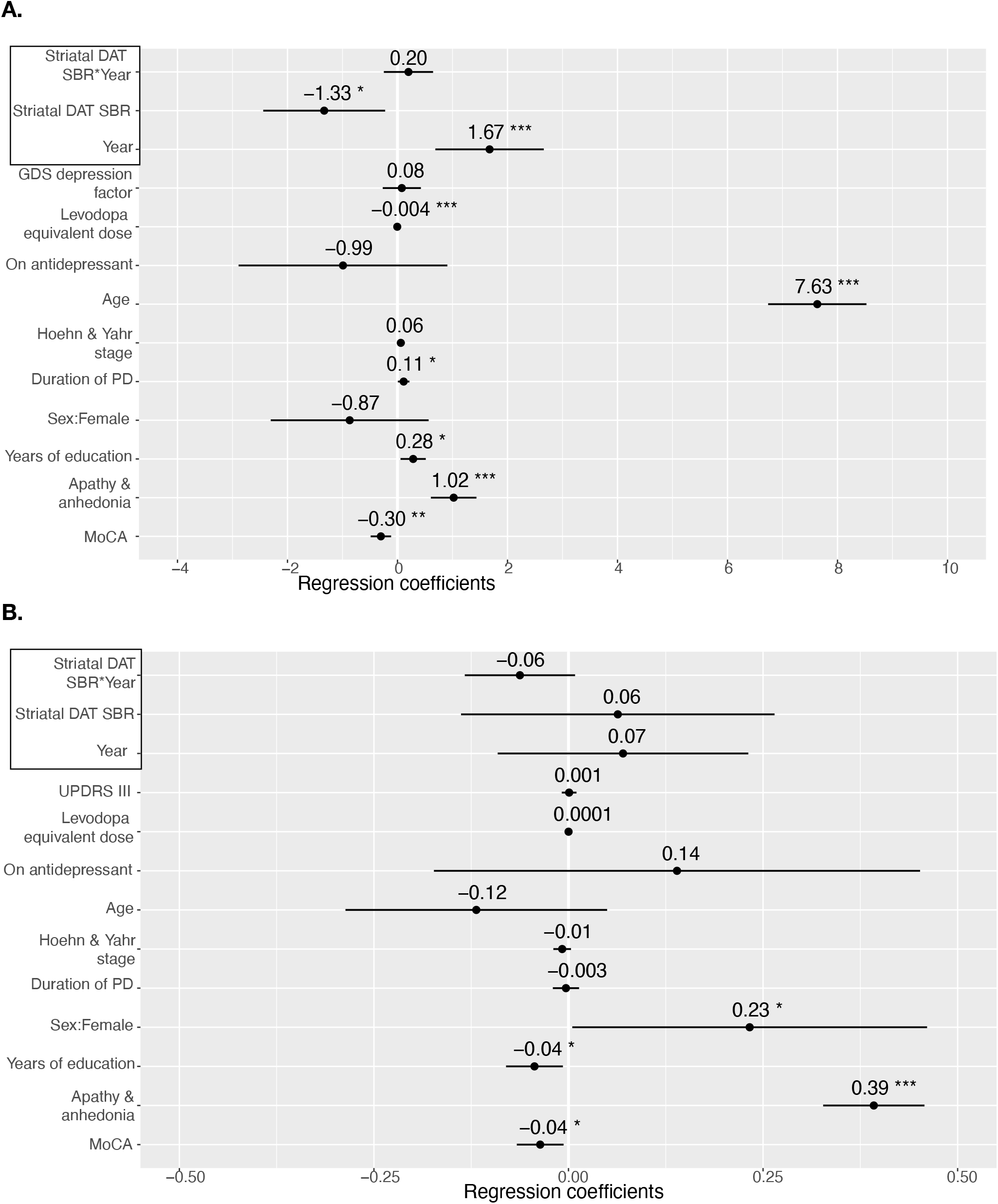
Sensitivity analyses examining the relationship between dopamine transporter (DAT) specific binding ratio (SBR) and motor and depression symptoms, using separate mixed-effects models. Apathy/anhedonia score has been replaced as the dependent variable with (**A**) MDS-UPDRS part III score and (**B**) the Geriatric Depression Scale-15 (GDS) “depression” factor. Points represent estimated regression coefficients and bars represent 95% confidence intervals; p<0.05*, p<0.01**, p<0.001***. The interaction between striatal dopamine transporter specific binding ratio (SBR) and time was non-significant for both models.

**Figure S2.**
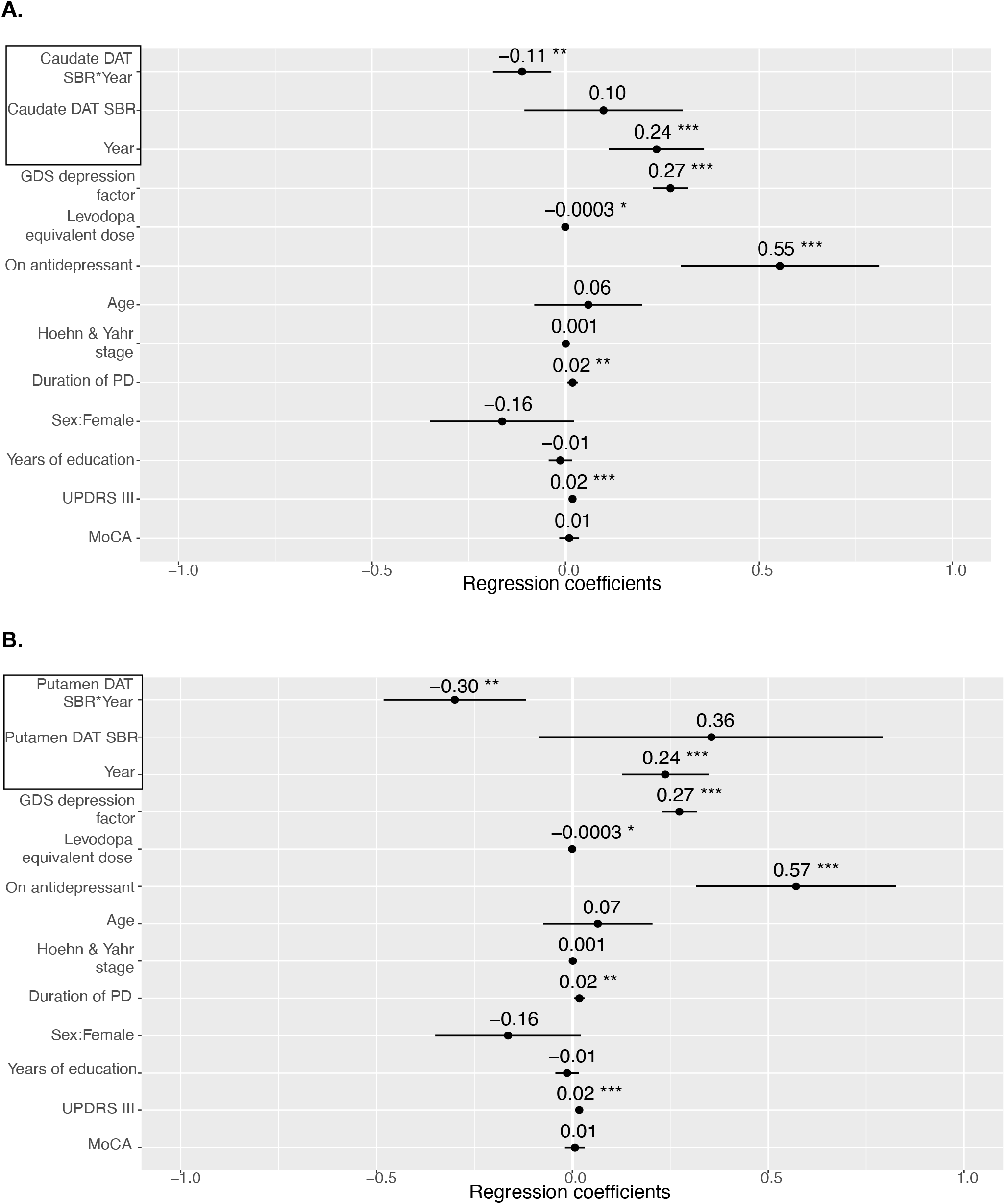
Mixed-effects model investigating the relationship between dopamine transporter (DAT) specific binding ratio (SBR) and apathy/anhedonia in different striatal subdivisions longitudinally. **A**. Caudate. **B**. Putamen. Points represent estimated regression coefficients and bars represent 95% confidence intervals; p<0.05*, p<0.01**, p<0.001***. Analysis of the association between apathy/anhedonia and DAT SBR in different striatal subdivisions found similar interactions with time in both the caudate ((β=-0.11, 95%CI [-0.19 -0.04], p<0.001) and putamen (β=-0.30, 95%CI [-0.48 -0.12], p<0.001; Figure S2).

**Figure S3.**
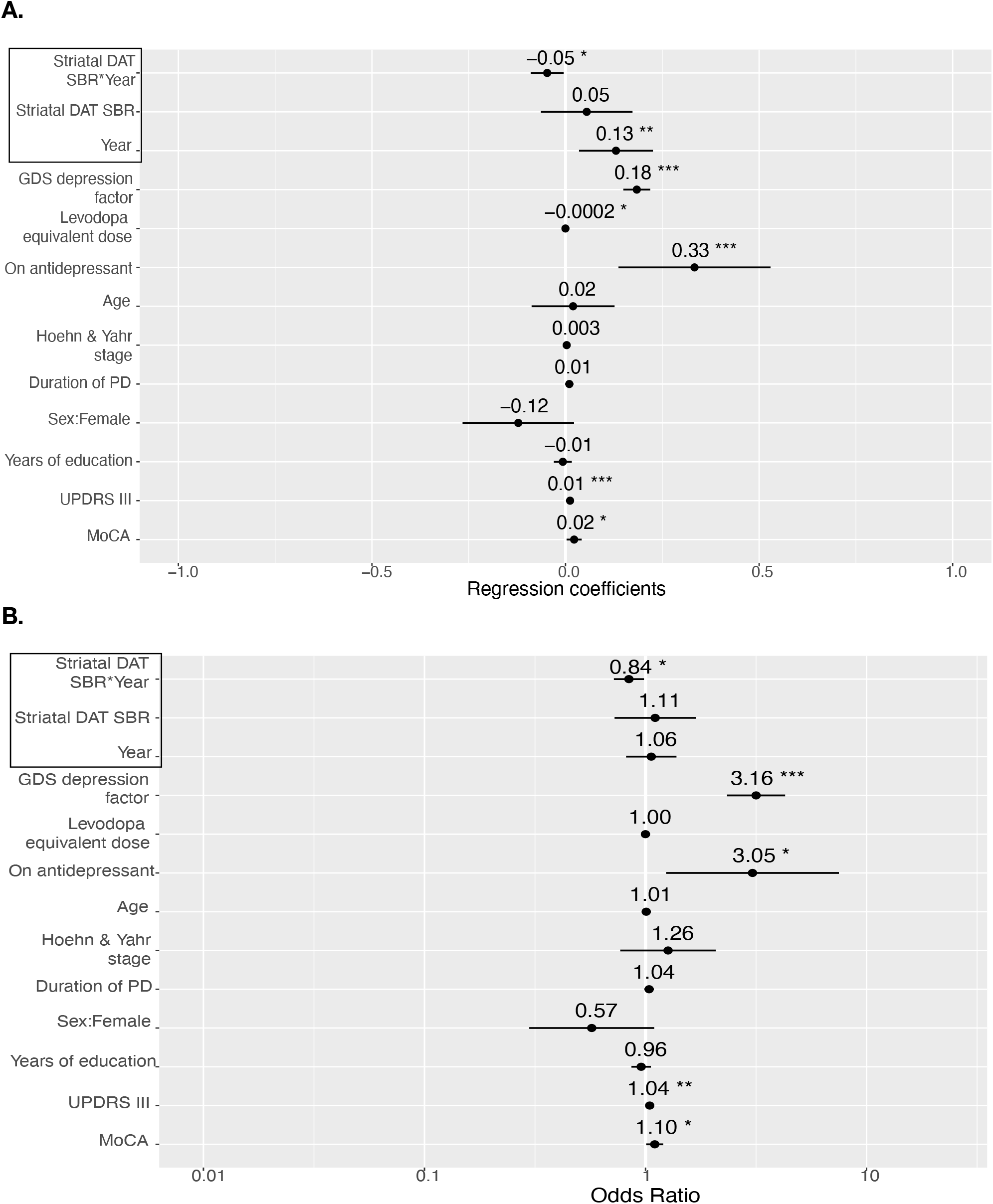
**A. Linear mixed effects model** investigating the relationship between striatal dopamine transporter (DAT) specific binding ratio (SBR) and the Geriatric Depression Scale (GDS)-3A, a three-item subset of the GDS-15, previously validated as a clinical measure of apathy. **B. Logistic mixed effects model** investigating the relationship between striatal DAT SBR and a categorical outcome using a cut-off of ≥2 on the GDS-3A that has high specificity for clinically relevant apathy. Consistent with our primary analyses, there was a significant interaction between striatal DAT SBR and time (Figure S3A) when treating the GDS-3A as a continuous outcome. Logistic mixed-effects modelling using a categorical GDS-3A dependent variable (score <2 or ≥2, indicating clinically significant apathy), revealed significantly increased odds of clinically relevant apathy with decreasing striatal DAT SBR as time progressed (odds ratio=0.84, 95%CI [0.73 0.98], p=0.031) (Figure S3B).

**Figure S4.**
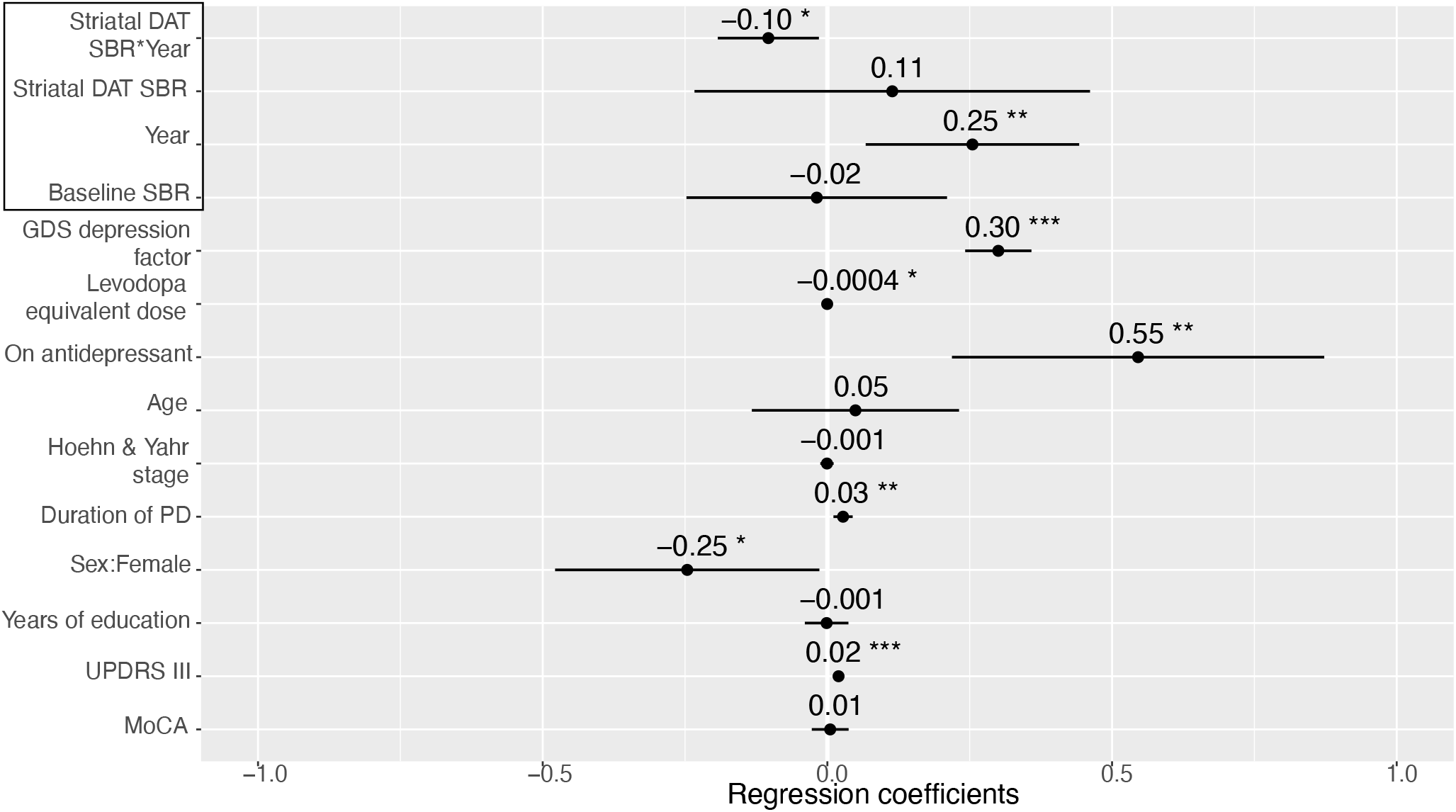
Mixed-effects model investigating the longitudinal relationship between striatal dopamine transporter (DAT) binding ratio (SBR) and apathy/anhedonia at post-baseline assessments, adjusted for baseline striatal DAT SBR (NB this makes the striatal DAT SBR main effect equivalent to change from baseline). Points represent estimated regression coefficients and bars represent 95% confidence intervals; p<0.05*, p<0.01**, p<0.001***. Baseline DAT SBR did not significantly predict subsequent apathy/anhedonia score (β=-0.02, 95%CI [-0.25 0.21], p=0.9), and the interaction between striatal DAT SBR and time remained significant (β=-0.10, 95%CI [-0.19 -0.01], p=0.023) (Figure S4).

